# Reusable Generic Clinical Decision Support System Module for Immunization Recommendations in Resource-Constraint Settings

**DOI:** 10.1101/2024.09.22.24314152

**Authors:** Samuil Orlioglu, Akash Shanmugan Boobalan, Kojo Abanyie, Richard D. Boyce, Hua Min, Yang Gong, Dean F. Sittig, Paul Biondich, Adam Wright, Christian Nøhr, Timothy Law, David Robinson, Arild Faxvaag, Nina Hubig, Ronald Gimbel, Lior Rennert, Xia Jing

## Abstract

Clinical decision support systems (CDSS) are routinely employed in clinical settings to improve quality of care, ensure patient safety, and deliver consistent medical care. However, rule-based CDSS, currently available, do not feature reusable rules. In this study, we present CDSS with reusable rules. Our solution includes a common CDSS module, electronic medical record (EMR) specific adapters, CDSS rules written in the clinical quality language (CQL) (derived from CDC immunization recommendations), and patient records in fast healthcare interoperability resources (FHIR) format. The proposed CDSS is entirely browser-based and reachable within the user’s EMR interface at the client-side. This helps to avoid the transmission of patient data and privacy breaches. Additionally, we propose to provide means of managing and maintaining CDSS rules to allow the end users to modify them independently. Successful implementation and deployment were achieved in OpenMRS and OpenEMR during initial testing.

## Introduction

The clinical decision support system (CDSS) is an effective tool^1,2^ to manage the clinical needs of patients. Rule-based CDSS is an important category for clinical practices that may account for 69% to 100% of the total CDSS usage in primary care settings in the USA^3^. However, these CDSS may become irrelevant to clinical practices if the associated rules are not kept up-to-date. Managing and maintaining CDSS rules can be resource-intensive, even for large academic medical centers, that are typically richer in resources than small medical practices.

Our group leverages technologies based on interoperability to develop reusable and sharable CDSS rules^4–6^; in addition, we implement and deploy the rules through two open-source electronic medical record (EMR) systems: OpenMRS and OpenEMR.

The former was first released in 2004^7,8^ and it does not have a CDSS module. It is the most commonly used open-source EMR system in the fields of clinical care, research, and development. Its current users are spread across 6,745 sites in 40 countries with 15.8 million active patients; many of these users work in resource-limited situations^7,8^.

OpenEMR is an open-source EMR and practice management system with over 4,000 downloads per month since its inception^9^; it is available in 30 languages and currently includes a CDSS module. However, it does not offer management, maintenance, and monitoring of the CDSS rules. Although CDSS can effectively provide consistent preventive clinical services^1,10^, its relevance and usefulness can become questionable if associated rules are not updated.

It is recognized that CDSS rule management and maintenance are very resource-intensive^11–14^. Therefore, sustainably introducing CDSS that is adapted to have reusable and updated rules, particularly in resource-limited settings, can benefit users immensely (primarily health care providers). Such an improvement in the CDSS will eventually benefit the patient populations that medical systems serve.

In this study, we demonstrate the feasibility of implementing reusable CDSS rules; as an example, (the CDC-recommended vaccination schedules^15^ are used to develop CDSS rules) in the two EMR systems. The aim of our work is to make the implementation process open-access, reusable, interoperable, and reproducible to enable the end users to manage and maintain CDSS rules independently. In this manuscript, we share the work currently in progress to achieve these aims, as well as the next steps.

## Methods

We leverage a couple of observed commonalities between EMR systems to develop our solution. OpenMRS^8^ and OpenEMR^9^ both have user interfaces that are displayed within a modern web browser and both EMRs support conversion of patient data to and from the fast healthcare interoperability resources (FHIR)^16^ format and the internal data model.

### Overall architecture

Figure 1. shows the overall architecture of the proposed solution to lack of reusable rules in CDSS. We use CDC immunization recommendations^15^ to create CDSS rules in the clinical quality language (CQL) format. These rules can be made publicly available in a repository as global rules that are downloadable by users for the creation of a local set of these rules that can be deployed on a case-by-case basis.

**Figure 1.**
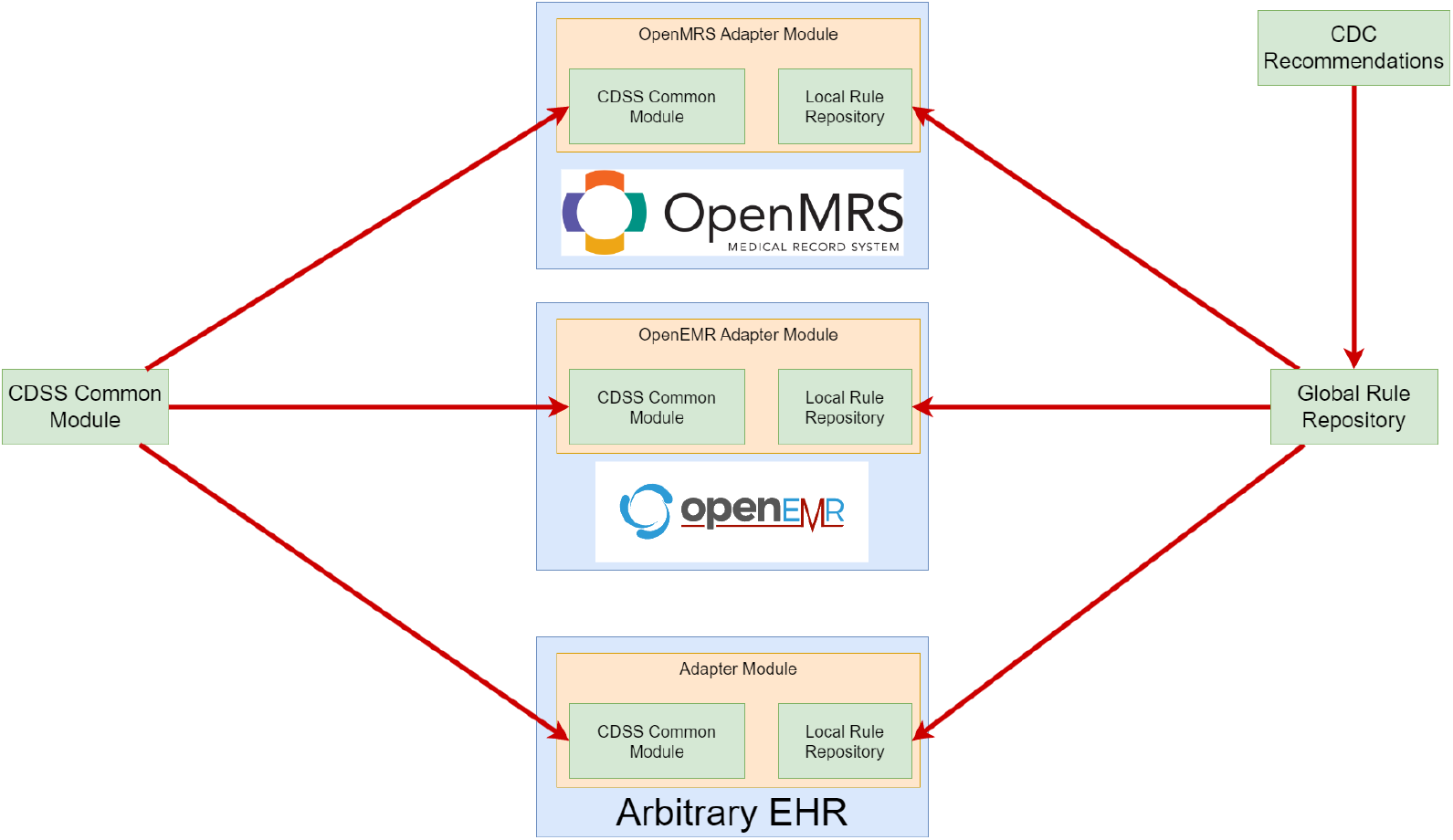
High-level architecture of the CDSS with a common-module to facilitate rule-sharing and customization across different platforms. The common-module interfaces with adapter modules for EMR systems, including OpenMRS and OpenEMR. Each adapter module includes a local rule repository, that can pulls rule recommendations from a global rule repository and provides individual customized recommendations for each patient.

In our proposed solution, the CDS engine is set into the client-side, where it runs in the web browser. This allows the CDSS engine to process data provided by the EHR in the fast healthcare interoperability resources (FHIR)^16^ format to calculate patient recommendations. The logic behind the decisions from the CDSS is not coded into the engine; instead, it originates from the CQL rules. The operating-system and technological stack of the EHR is irrelevant to the CDSS engine. We call this CDSS engine the “common-module” as it is designed to work universally, despite technological inconsistencies.

Despite the uniformity of the common-module for available EMRs, specific functionalities must be added to make it better compatible with individual EMR and operate optimally. For this purpose, we introduce the adapter module. The adapter module is tailored for a given EMR to properly integrate it with the common-module and provide persistent storage for rules, and recommendation display on user interfaces. The adapter module also supplies the common-module with all the resources needed for the proper execution of rules.

In ongoing studies, we aim to improve the rule-based CDSS solution proposed here with additional features. For instance, in addition to executing the rules for patient-specific vaccine recommendations, the module will also track past CDSS rules usage to provide data-driven evidence for future optimization and will enable an individual or organization to modify the rules according to their requirements.

### Creation of rules

For rule-based CDSS, CDSS rules added to the CDSS in EMRs are logical components to determine patient-specific recommendations, given the patient’s clinical history. Currently, rules are written manually in CQL^17^ and utilize FHIR v4.0.1^16^ representations of objects to compute and generate recommendations. Figure 2. shows an example of a rule for a recommendation regarding the measles, mumps, and rubella (MMR) vaccine.

**Figure 2.**
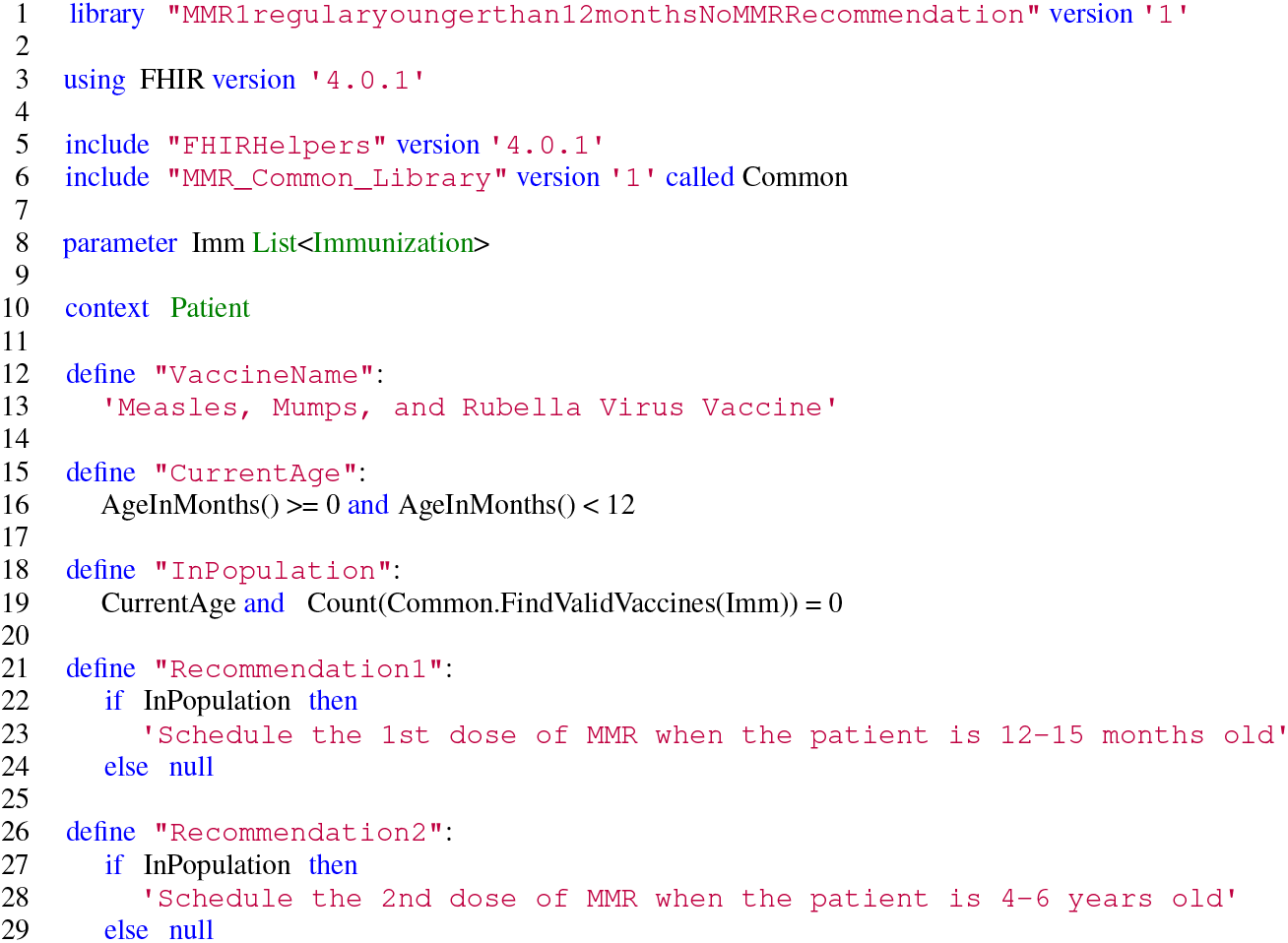
Example of a sample CQL rule for the measles, mumps, and rubella (MMR) vaccine that checks if the patient is between 0 and 12 months old and if they have not been administered a prior MMR vaccination. If so, it is recommended that the clinician should *‘Schedule the 1st dose of MMR when the patient is 12-15 months old’* and *‘Schedule the 2nd dose of MMR when the patient is 4-6 years old’*

We organize CDSS rules by vaccine and follow a few principles to create them:

- Parameters are a list of FHIR^16^ resources, such as all immunizations administered to a patient or all clinical observations of the patient. Parameters represent all FHIR^16^ resources of a single type in a patient’s record. The logic in CQL files filters the relevant resources of interest from the list for processing in the rule. In Figure 2, the CDSS rule uses the parameter *Imm* on line 8, which is a list of immunizations.
- Rules must have at least one expression called “Recommendations” or “Recommendation” followed by an integer. This is what will be displayed to the clinician as shown in lines 21 and 26 in Figure 2.
- We utilize libraries to store useful functions and value sets for the rules, which allows the rule to be readable and concise. In Figure 2. the CDSS rule utilizes the *MMR_Common_Library* on line 6, demonstrating this point.

Rules formatted in CQL must be converted into expression logical model (JSON-ELM) before they can be deployed in the common-module. This conversion can be completed automatically using the cql-to-elm-cli^17^ tool or by a Docker service called the cql-translation-service^18^.

### Rule testing

To verify that CDSS rules are syntactically and logically correct, we utilize the cdss-testing-harness^19^, which is a modified version of cql-testing-harness^20^. It is a framework for unit testing of CQL^17^ files within a Node.Js^21^ environment.

The cdss-testing-harness^19^ automatically converts CQL files into the JSON-ELM by creating a temporary Docker container^18^. For this study, we created sample patients, immunization records and observations in the FHIR format to test all possible logical branches of the rules. The test cases were stored in a directory hierarchy. We also wrote the unit tests for every rule, to ensure rule correctness for positive and negative cases. The test cases were incorporated into the unit tests by the cdss-testing-harness^19^. Once rules were thoroughly tested, they could be applied in the CDSS.

### The common-module

The key component in this project is the common-module which is written in Javascript and embedded directly into the webpages of the EMR. It enables the execution of JSON-ELM rules, solely on the client-side. The common-module depends on cql-execution^22^ for executing the logic within the rules, cql-exec-fhir^23^ to process FHIR^16^ data types and a modified cql-exec-vsac^24^ called browserfy-exec-vsac^25^ to resolve the value sets.

The task of the common-module is to gather the FHIR^16^ resources and the value sets needed to execute the rules. To do this, the common-module contains several utility functions to request FHIR^16^ resources, convert different data types, and other functionalities. The process of executing a rule within the common-module is shown in Figure 3. The key to the flexibility of the common-module is the endpoints-map, which is a flexible and adaptable data structure that “describes” how to retrieve resources from the EMR.

**Figure 3.**
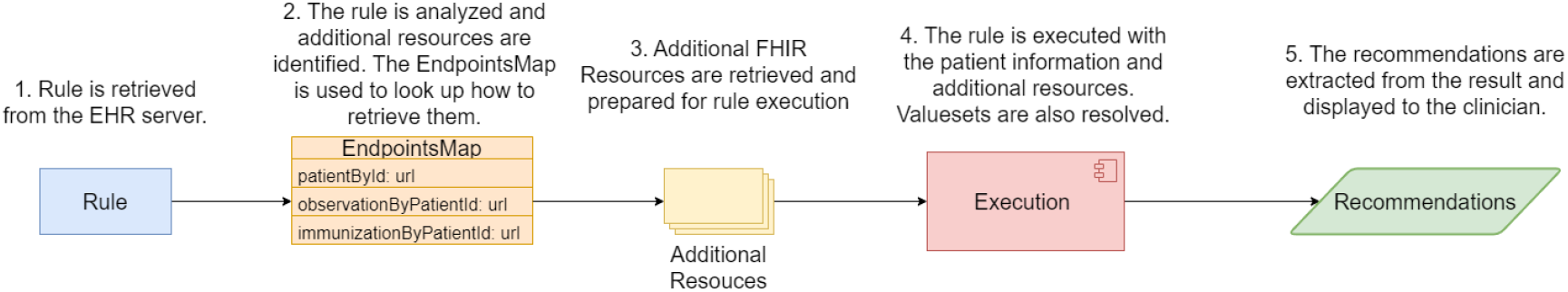
The process of executing rules within the common-module.

For easy distribution, the entire common-module is bundled into a single Javascript file with Webpack^26^.

*Endpoints-Map* The endpoints-map is a key-value lookup table describing the method of retrieving the resources. The endpoints-map is configured on a per-instance basis, using the specifics of the EMR. When the common-module needs a resource, it refers to the endpoints-map to find out how to retrieve the resource. This can be done either by HTTP request (as in the case of FHIR^16^ resources) or a function if the request cannot be a simple HTTP request. Generally, the endpoints-map is configured specifically for the EMR that the CDSS is deployed in, therefore, it allows the system to flexibly adopt to different configurations. Figure 4. shows an endpoints-map that is configured to request resources from a hypothetical EMR.

**Figure 4.**
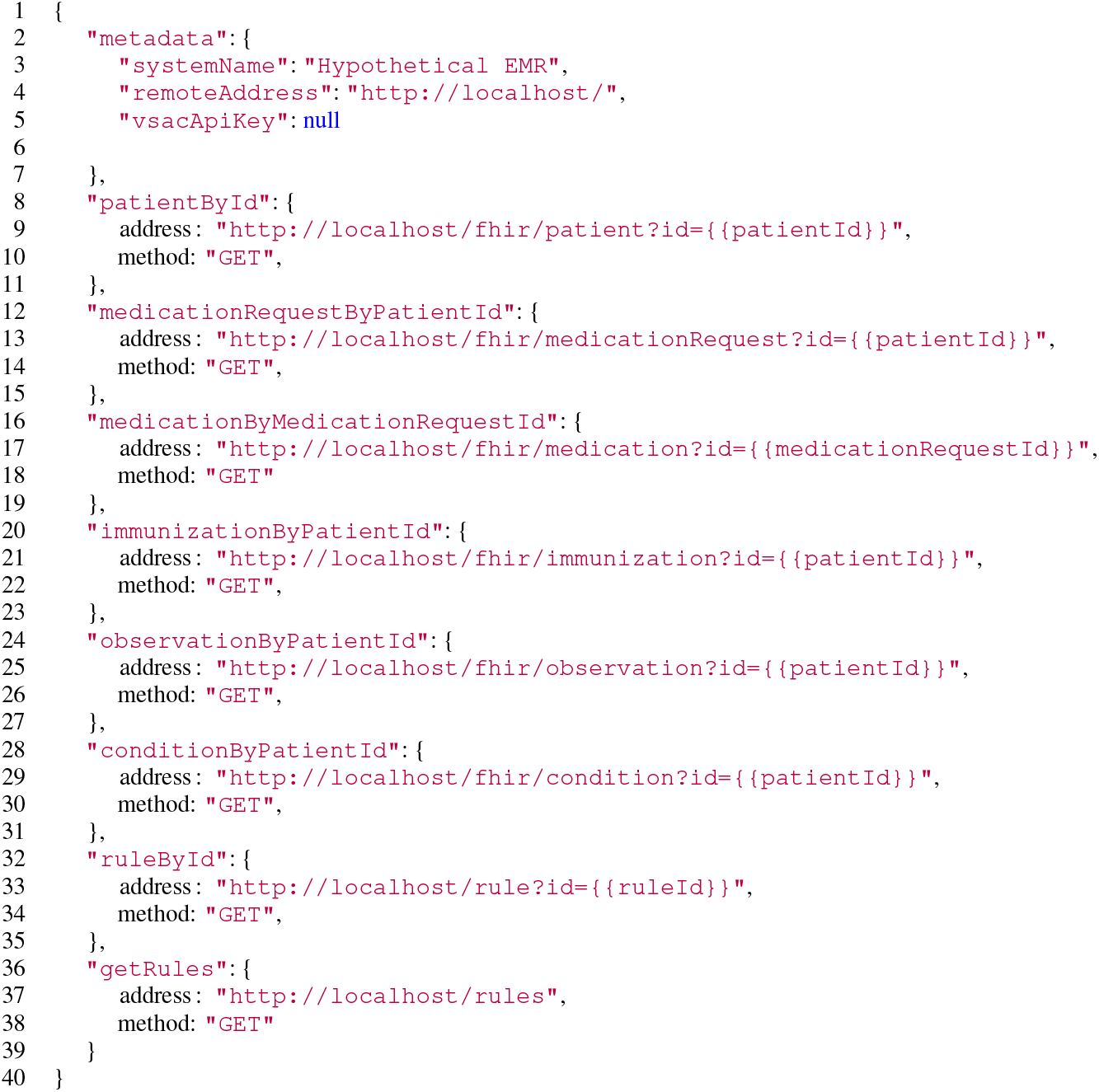
A sample configuration of the endpoints-map for a hypothetical EMR running on localhost. The section on *metadata* provides system-level details. The other sections define the endpoints that should be used to retrieve FHIR resources, such as patients, conditions, observations and immunizations. The URLs also use placeholder parameters, such as *{{patientId}}*, for dynamic queries. The rules section define which URLs must be used to obtain JSON-ELM forms of the rules.

*Browserfy-cql-exec-vsac* The common-module requires a method to resolve the value sets declared in the rules. For this purpose, a package called cql-exec-vsac^24^ is typically used. However, cql-exec-vsac^24^ works on the server-side. To work around this limitation, we adapted the original cql-exec-vsac^24^ to create a client-side version called browserfy-cql-exec-vsac^25^, which has the same functionality as the cql-exec-vsac^24^, but it does not provide system storage and offers customizable configuration of the value set server.

### Incorporating the common-module into the EMR

Many EMRs including OpenMRS and OpenEMR can be to customized to have additional functionalities by incorporation of suitable modules. These modules have direct access to EMR and utilize its software development kit (SDK). Similarly, the common-module must also be integrated and embedded into the web pages of the EMR. This process is facilitated by introducing an adapter module, which is simply an EMR module to properly execute the common-module.

The adapter module has a few responsibilities; its main responsibility is to embed the common-module into the web pages of the EMR and to configure the endpoints-map to the exact specifications of the EMR. Considering that every EMR is different, the adapter module must configure the endpoints-map in the common-module accordingly.

The adapter modules have direct access to the EMR resources, and thus, they can serve additional functions. For example, the adapter module can persistently store rules in an EMR; additionally, it can provide an application programming interface (API) to retrieve rules as needed by the common-module. Last but not least, the adapter module can also facilitate the tracking of rule usage by saving every instance when a rule is triggered in an internal EMR database.

### Rule management

We are currently developing the ability for qualified users to modify rules within their local EMR environment through its user interface. However, any changes to the rules must be tested, and the rules in CQL must be converted to JSON-ELM before implementation in the CDSS. For this, our ongoing work involves building a web service that automatically accepts changes in rules, tests them with the aforementioned rule testing environment, and upon success, sends the changed rules back to the JSON-ELM for implementation in the CDSS.

## Results

### Common-module

The common module was completed and successfully tested in both OpenMRS and OpenEMR. Ongoing work, expected to be released in the future, focuses on the maintenance of the common module and improvements to it.

### OpenMRS

OpenMRS^8^ is designed in a modular manner, where the modules implement the functionality of the software. The OpenMRS 3 modules have two layers: the back-end and front-end. The back-end is Java^27^ based, that utilizes the Spring^28^ framework with MySQL^29^ or MariaDB^30^ as databases. The front-end is built with React^31^ and Typescript^32^.

The adapter module for OpenMRS consists of two parts: the Java^27^ based back-end called *cdss*^33^ to create API endpoints, add tables to the database, and configure some OpenMRS settings. The front-end module called *openmrs-esm-cdss-app*^33^ allows the user to interact with the *cdss*^33^ system. It adds a new section to the patient chart view (Figure 5) and new pages to view all the events when the CDSS system was used.

**Figure 5.**
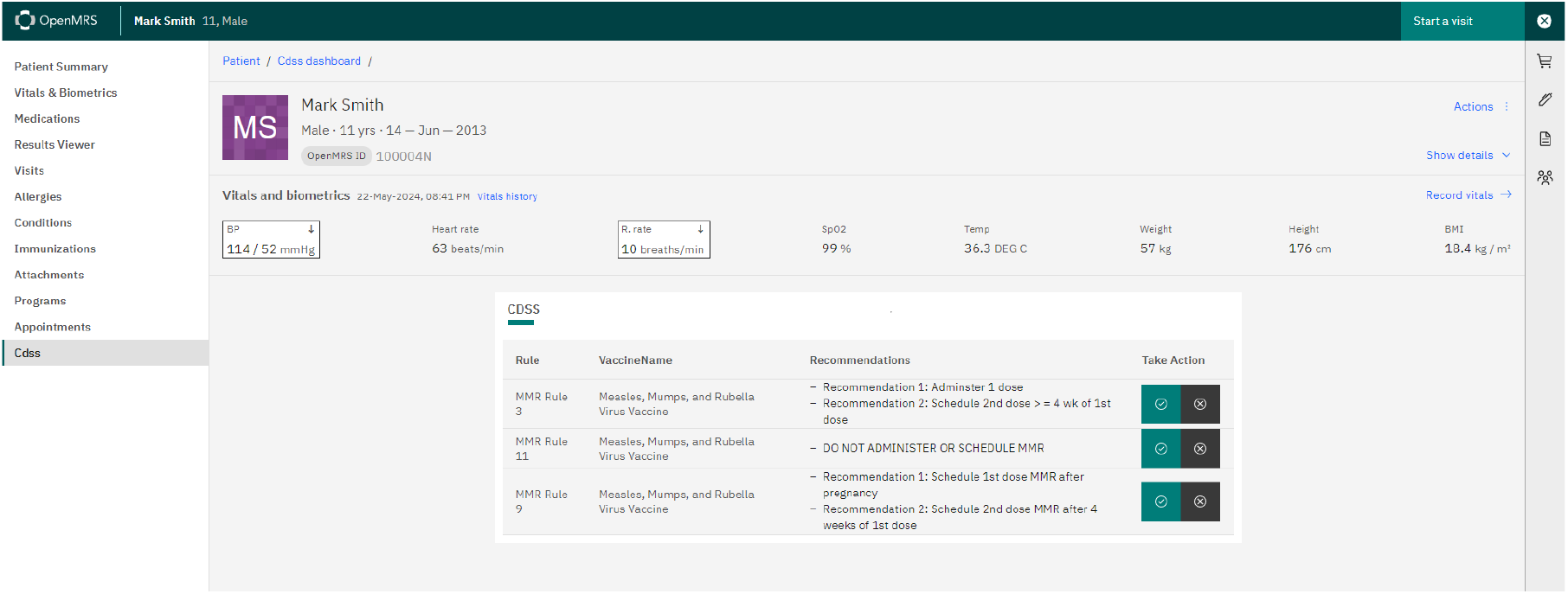
Patient chart in OpenMRS coupled to the CDSS for hypothetical patient. Recommendations are shown in the 3rd column of the table.

**Figure 6.**
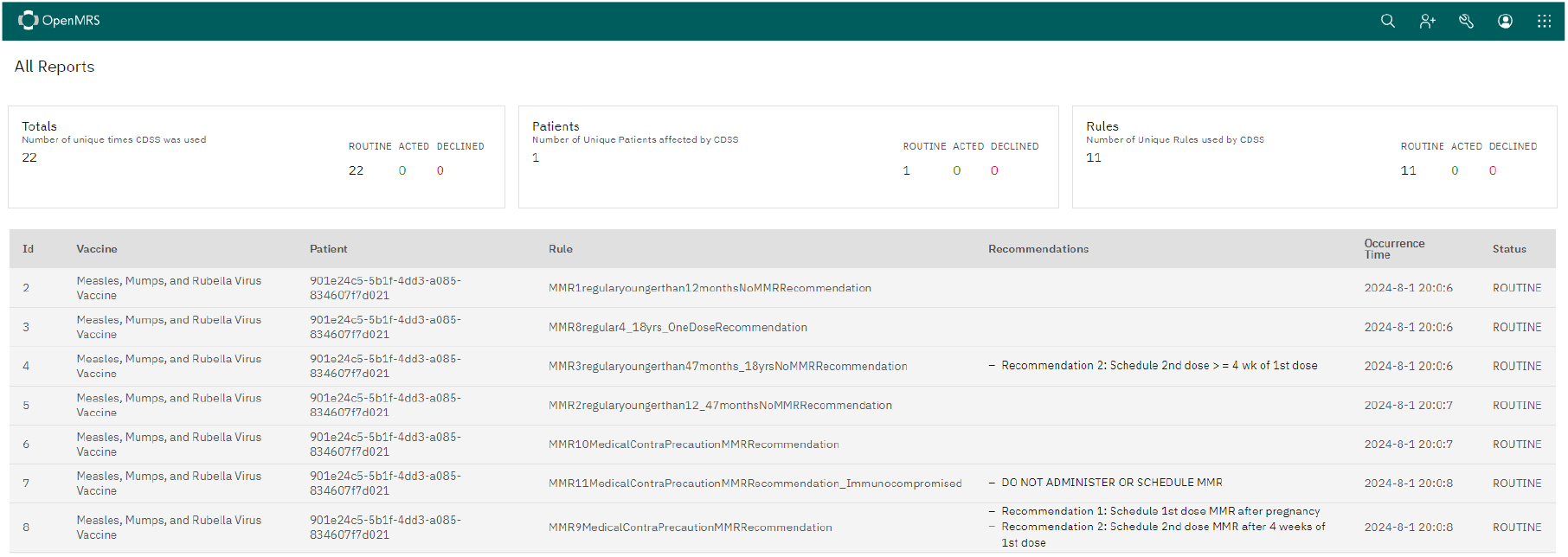
The page in OpenMRS where one can view all the past consultation of the CDSS. The entries on this page are displayed whether the system passes a recommendation or not.

### OpenEMR

OpenEMR’s back-end relies on MySQL^29^ or MariaDB^30^ as the database management system to store patient information and related data. The logic on the server-side is primarily written in PHP^34^, that interacts with the database. The integration of health information systems and standards, such as FHIR^16^, for data exchange is supported by OpenEMR. Configuration is managed through PHP^34^ configuration files, which include setting up the environment variables and server connections. OpenEMR is very customizable software that can be easily adapted to various healthcare settings.

The adapter module developed for OpenEMR^9^ was built using the custom module framework provided by OpenEMR^9^; this adapter module was integrated with the CDSS with a webpacked^26^ common-module^33^. As shown in Figure 7, a large recommendation tile is provided on the patient dashboard in the OpenEMR^9^ interface by the adapter module, and the patient’s FHIR^16^ data is processed according to the rules. The recommendation tile displays the results, which may include clinical insights or recommendations based on the patient data.

**Figure 7.**
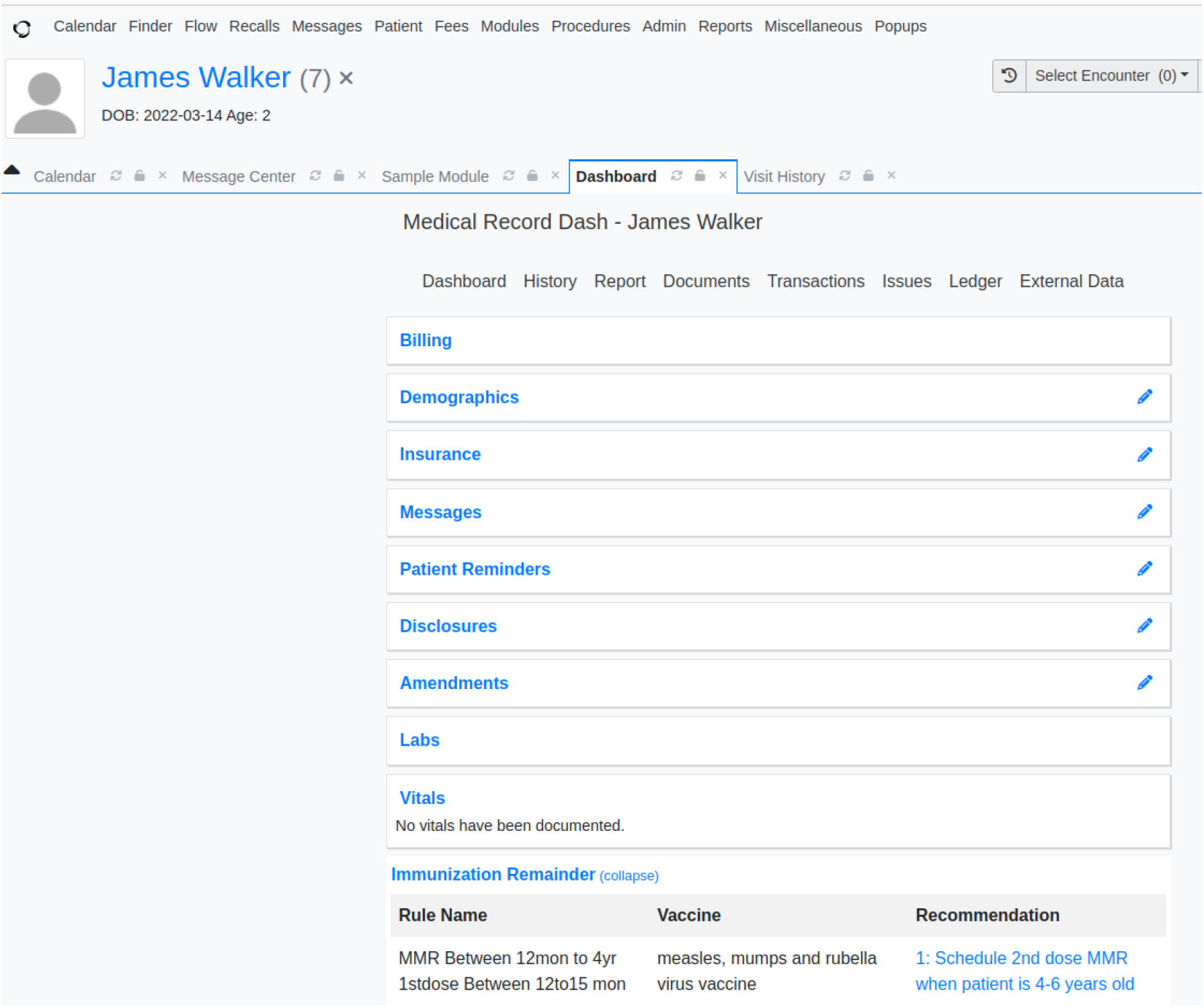
Patient chart for a hypothetical patient in OpenEMR coupled to the CDSS. Recommendations are shown in the Immunization Reminder section.

### CDSS Rules

The project on reusable rules is still actively being developed, despite the promising results presented in this report. Certain features have been completed, and others are in progress. Currently, we have completed and validated 13 categories, 19 vaccines, and 465 rules in tabular and chart formats. We have also completed and tested 12 CQL rules, while the rest are being developed and assessed.

## Discussion

In this study, we investigate the deployment of a CDSS module with multiple EMRs to assess the potential of standardizing the method and generating good output, i.e., patient-specific recommendations, from multiple platforms. The CDSS developed here is EMR-agnostic. It needs only a browser-based user interface and FHIR^16^ resources from the EMR to function properly. In addition, as the CDSS runs on the client-side, there is no need to set up additional services or to transfer patient data outside the institution, minimizing the risk of violating data privacy.

However, these benefits come at the cost of efficiency. The CDSS runs only on the client-side, which is considered a benefit; on the downside, this configuration causes the CDSS to be heavily reliant on the processing speed of the client-side device. Future research should focus on testing the performance and optimizing the common-module to enhance CDSS efficiency.

It is important to mention that CDS Hooks^35^ offers the same functionality as our system; it invokes CDSS through the workflow of the clinician. However, CDS Hooks^35^ and the CDSS module developed here are implemented differently, where CDS Hooks^35^ requires a CDS server to determine the recommendations. In resource-constrained settings, acquiring additional services for CDS may not be feasible. Contrarily, the CDSS module does not require additional hardware, and thus, it offers a unique value for resource-limited situations.

Last but not least, the CDSS module developed here is based on Javascript and runs on the client-side only. Therefore, it can feasibly be structured as a SmartOnFHIR^36^ app to encourage its adoption among users and institutions. The SmartOnFHIR^36^ app is out of the scope in our this study, but this is an important area of development to adapt the CDSS proposed here into a SmartOnFHIR^36^ app in the future.

## Conclusion

In this study, we demonstrate that the same CDSS module can be deployed on disparate and technological dissimilar EMR systems, such as the OpenEMR and OpenMRS, to generate consistent patient-specific recommendations. Importantly, we demonstrate that it is possible to achieve a CDSS system with reusable rules without setting up additional services. After thorough testing, we will make the codes, documentation, and CDSS rules publicly available. We anticipate to provide additional resources for communities with limited resources to use in clinical practices. The CDSS modules and rules can also be used in education or research to avoid duplicate efforts.

## Data Availability

All data produced in the present study are available upon reasonable request to the authors and all the codes will be available through GitHub repository after thorough testing.

## Acknowledgments

This study is made possible by funding from the National Institute of General Medical Sciences (R01GM138589) and the Office of Data Science Strategy (3-R01-GM138589-03S1) at the National Institutes of Health, with additional support from grants P20GM121342 and T15LM013977.

